# Vaccine effectiveness against hospitalisation and comparative odds of hospital admission and severe outcomes with BQ.1, CH.1.1. and XBB.1.5 in England

**DOI:** 10.1101/2023.07.28.23293333

**Authors:** Freja C M Kirsebom, Katie Harman, Rachel J Lunt, Nick Andrews, Natalie Groves, Nurin Abdul Aziz, Russell Hope, Julia Stowe, Meera Chand, Mary Ramsay, Gavin Dabrera, Meaghan Kall, Jamie Lopez Bernal

## Abstract

**Background:** Since the first emergence of Omicron BA.1 in England in November 2021, numerous sub-lineages have evolved. In September 2022, BA.5 dominated. The prevalence of BQ.1 increased from October, while the prevalence of CH.1.1 and XBB.1.5 increased from December 2022 and January 2023, respectively. Little is known about the effectiveness of the vaccines against hospitalisation with these sub-lineages, nor the relative severity.

**Methods:** A test-negative case-control study was used to estimate the incremental effectiveness of the bivalent BA.1 booster vaccines against hospitalisation, relative to those with waned immunity where the last dose was at least 6 months prior. The odds of hospital admission for those testing PCR positive on the day of an attendance to accident and emergency departments and the odds of intensive care unit admission or death amongst COVID-19 admissions were compared between variants. Additionally, a Cox proportional hazards survival regression was used to investigate length of stay amongst hospitalised cases by variant.

**Findings:** There was no difference in incremental vaccine effectiveness against hospitalisation with BQ.1, CH.1.1 or XBB.1.5, nor was there a difference in the severity of these variants. Effectiveness against hospitalisation was 48.0% (95% C.I.; 38.5-56.0%), 29.7% (95% C.I.; 7.5-46.6%) and 52.7% (95% C.I.; 24.6-70.4%) against BQ.1, CH.1.1 and XBB.1.5, respectively, at 5 to 9 weeks post booster vaccination. Compared to BQ.1, the odds of hospital admission were 0.87 (95% C.I.; 0.77-0.99) and 0.88 (95% C.I.; 0.75-1.02) for CH.1.1 and XBB.1.5 cases attending accident and emergency departments, respectively. There was no significant difference in the odds of admission to intensive care units or death for those with CH.1.1 (OR 0.96, 95% C.I.; 0.71–1.30) or XBB.1.5 (OR 0.67, 95% C.I.; 0.44-1.02) compared to BQ.1. There was also no significant difference in the length of hospital stay by variant.

**Interpretation:** Together, these results provide reassuring evidence that the bivalent BA.1 booster vaccines provide similar protection against hospitalisation with BQ.1, CH.1.1 and XBB.1.5, and that the emergent CH.1.1 and XBB.1.5 sub-lineages do not cause more severe disease than BQ.1.

**Funding:** None.

## Introduction

The first Omicron sub-lineage to emerge in the UK was BA.1 in November 2021^1^, followed by BA.2^2^ and BA.4 and BA.5^3^. In the autumn/winter of 2022/23, BA.5 dominated in September. The prevalence of BQ.1/BQ.1.1 increased from October^4^, while the prevalence of CH.1.1 and XBB.1.5 increased from December 2022 and January 2023, respectively^5^ (Supplementary Figure 1). These sub-lineages have all acquired different combinations of mutations in the spike protein as compared to BA.1^6,7^. Both CH.1.1 and XBB.1.5 have demonstrated growth advantages and proven to be highly transmissible sub-lineages of Omicron^7^.

Previous Omicron sub-lineages have shown no increase in severity, including BA.4 and BA.5 compared to BA.2, and BA.4.6, BA.2.75 and BQ.1 compared to BA.5^8,9^, but there is limited evidence available on the severity of CH.1.1 and XBB.1.5. Initial data suggested XBB.1.5 has a similar level of severity compared to the baseline of BQ.1^10^. Evidence from laboratory-based assessments of the efficacy of therapeutic monoclonal antibodies against BQ.1, BA.2.75.2 (parental lineage of CH.1.1) and XBB (parental lineage of XBB.1.5), as well as studies evaluating the neutralising ability of plasma antibodies from vaccinated individuals against these variants have suggested significant immune escape as compared to that observed against the wild-type, BA.1 and BA.5 strains^6,7,11–15^. However, neutralising assays for previous sub-lineages have often shown reduced neutralising which has not translated to a reduction in the real-world effectiveness against severe disease outcomes^16,17^. To our knowledge, there are no real-world estimates of VE against BQ.1 or CH.1.1. Studies from the United States (US) found VE against infection and hospitalisation with XBB/XBB.1.5 was generally comparable to that seen against BA.5^18,19^, while a study from Singapore found protection against A&E attendance was 49% during an XBB wave^20^.

As part of the UK COVID-19 vaccination programme, an autumn 2022 booster programme commencing 5th September 2022 was recommended by the JCVI and bivalent BA.1 boosters with either Pfizer BioNTech (Original/Omicron BA.1 Comirnaty®) or Moderna (Spikevax® bivalent Original/Omicron BA.1 vaccine) were offered to all adults aged 50 years and over and vulnerable individuals, including the immunosuppressed^21,22^.

Here, we use national-level electronic health records from England to estimate the incremental vaccine effectiveness (iVE), often also called relative VE^23,24^, of the bivalent BA.1 boosters against hospitalisation with BQ.1, CH.1.1. and XBB.1.5 in England. We also assess the relative severity of these variants by estimating the odds of hospital admission or death following accident and emergency (A&E) attendance, for both CH.1.1 and XBB.1.5 compared to BQ.1. As a secondary indicator of variant severity, we estimate the odds of intensive care unit (ICU) admission or death amongst hospitalised cases by variant.

## Methods

### Study design

To estimate VE of the bivalent BA.1 booster vaccines offered as part of the autumn 2022 booster programme against hospitalisation by variant, a TNCC study design was used where positive PCR tests from hospitalised individuals aged 50 years and older are cases while negative tests from such individuals are controls, as previously described^16,17,24–26^.

To estimate the odds of hospital admission by variant, individuals of any age with a positive PCR test attending A&E who went on to be admitted or transferred with a length of stay of 2 or more days, or whose attendance ended in death, or who died within 2 days of their A&E attendance were included, as well as comparable individuals who did not go on to be admitted, as previously described^8^.

To estimate the odds of ICU admission or death (referred to in this manuscript as severe outcomes following hospitalisation) amongst cases admitted to hospital, individuals aged 50 years and older who were hospitalised with COVID-19 and who were admitted to ICU or who died were included, as well as comparable individuals who did not require ICU and did not die. Additionally, a Cox proportional hazards survival regression was used to investigate length of stay amongst hospitalised cases by variant.

### Data sources

Full details of all data sources are available in the Supplementary Appendix. All data sources are national-level healthcare datasets which include the entire relevant population in England. The study period for tests contributing to all analyses was from 5^th^ December 2022 to 2^nd^ April 2023, when the variants of interest were co-circulating.

To estimate VE, hospital based positive and negative PCR tests were extracted, as previously described^16,17,24–26^. To estimate the odds of hospital admission and severe outcomes in hospital, only PCR positive individuals were included. Variant status was identified by whole genome sequencing information from the national variant line list, coordinated by the COVID-19 Genomics UK consortium^27^. Only individuals with BQ.1, CH.1.1 or XBB.1.5 were retained.

Data were linked to the National Immunisation Management System NIMS as previously described^16,17,24–26,28^ and accessed for dates of vaccination and manufacturer, sex, date of birth, date of death, ethnicity, and residential address. For VE analyses the following individuals were excluded: those who were unvaccinated, those who had received only one dose, trial doses, an autumn dose without receiving at least two other doses prior to 5^th^ September, an autumn booster less than 12 weeks after their next most recent dose, two autumn doses, a vaccine coded as bivalent prior to 5^th^ September, an autumn dose not coded as bivalent, and those whose last dose prior to 5^th^ September 2022 was by a manufacturer other than AstraZeneca, Pfizer or Moderna^24^.

To assess the odds of hospital admission following A&E attendance, cases were linked to the Emergency Care Data Set (ECDS) and Secondary Uses Service (SUS). Only those who attended A&E on the same day as their first positive test were included^29^. SUS data were used to identify subsequent hospital admissions where the length of stay was at least 2 days. SUS data was also used to estimate VE against hospitalisation and the odds of ICU or death amongst those hospitalised, regardless of ECDS admission status. Admissions were restricted to those with a date of test 1 day before up to 2 days after the admission date and where the length of stay was at least 2 days. ICD-10 codes from the primary diagnosis field were used to classify acute respiratory illness (ARI) (Supplementary Table 1). Classification of Interventions and Procedures (OPCS-4) codes were used to identify individuals who received treatment on intensive care unit (ICU)^25^.

### Covariates and adjustment

For all analyses, week of test date, gender, age group, region, IMD quintile and reinfection status were included as potential confounding variables. For VE and odds of ICU admission or death and length of stay analyses, ethnicity, risk group status, care home status and health and social care worker status were included as additional confounders. Vaccination status was an additional confounder for all severity analyses.

### Statistical analysis

To estimate VE, multivariable logistic regression was used with the test result as the outcome, vaccination status as the primary variable of interest and with confounder adjustment as described above. VE was calculated as 1- odds ratio and given as a percentage. Incremental VE of the bivalent booster was estimated amongst those who had received at least two doses prior to the 5^th^ September 2022 and whose final dose prior to the 5^th^ September 2022 was at least 6 months before their test date, with those who did not receive a bivalent booster being the comparator group. VE was estimated at the following intervals since booster vaccination; 0 to 6 days, 7 to 13 days, 2 to 4 weeks, 5 to 9 weeks, 10 to 14 weeks, or 15 or more weeks. Analyses were restricted to those aged 50 years and older and estimated by manufacturers combined. Sensitivity analyses were conducted to estimate iVE for those with ARI ICD-10 coding in the primary diagnosis field.

To assess the odds of admission or death following A&E attendance by variant, conditional logistic regression models were used to estimate odds ratios of the outcome for both CH.1.1 and XBB.1.5 compared to BQ.1. Models were stratified by test week and with confounder adjustment as described above.

To estimate the odds of ICU or death amongst hospitalised cases by variant, multivariable logistic regression with ICU admission or death as the outcome, variant as the primary variable of interest and with confounder adjustment as described above. Sensitivity analyses were conducted to estimate the odds of ICU admission or death for those with ARI ICD-10 coding in the primary diagnosis field. To estimate length of stay amongst hospitalised cases, a Cox proportional hazards survival regression was used. Variant was included as an independent variable with confounder adjustment as described above. Only individuals who had an admission date, discharge date and a length of stay between 0 and 21 days (to ensure all individuals in the study period had time to be discharged and to allow for delays in the SUS hospitalisation data reporting) were included. Individuals who died were excluded to avoid bias. Model outputs are reported as the predicted median length of stay.

## Results

### Vaccine effectiveness against hospitalisation

There were 191,229 eligible tests from hospitalised individuals aged 50 years and older, with 1,647 BQ.1 cases, 877 CH.1.1 cases, 1,357 XBB.1.5 cases and 187,348 test negative controls. Full descriptive characteristics are available in Supplementary Table 2 and Supplementary Figure 2.

The iVE of the bivalent BA.1 boosters was 48.0% (95% C.I.; 38.5-56.0%), 29.7% (95% C.I.; 7.5- 46.6%) and 52.7% (95% C.I.; 24.6-70.4%), in addition to the protection from previous doses, against BQ.1, CH.1.1 and XBB.1.5, respectively, at 5 to 9 weeks post vaccination. iVE against all sub-lineages waned over time, and iVE was 30.5% (95% C.I.; 18.7-40.6%), 24.5% (95% C.I.; 8.6-37.7%) and 21.1% (95% C.I.; 9.6-31.1%) against BQ.1, CH.1.1 and XBB.1.5, respectively, at 15 or more weeks post vaccination (Table 1, Figure 1). Point estimates were lower for CH.1.1 and XBB.1.5 than for BQ.1 at most time points, but confidence intervals overlapped, and this difference was not statistically significant. Sensitivity analyses found there was no difference in iVE when we restricted to only include hospitalisations with a respiratory code in the primary diagnosis field (Supplementary Table 3).

**Figure 1.**
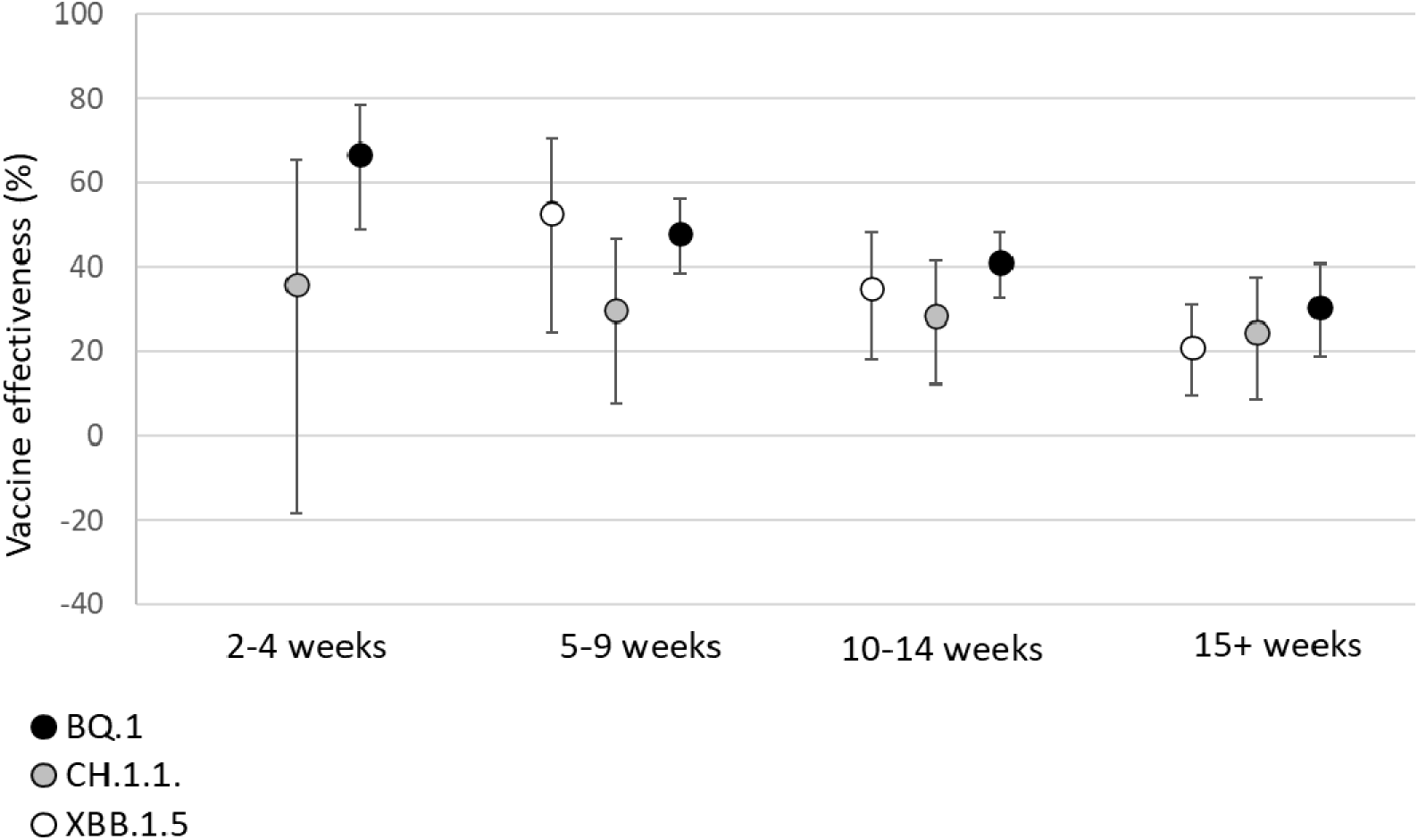
Incremental vaccine effectiveness (iVE) against hospitalisation of the bivalent BA.1 booster vaccine against BQ.1, CH.1.1 and XBB.1.5 amongst adults aged 50 years and older in England.

**Table 1.**
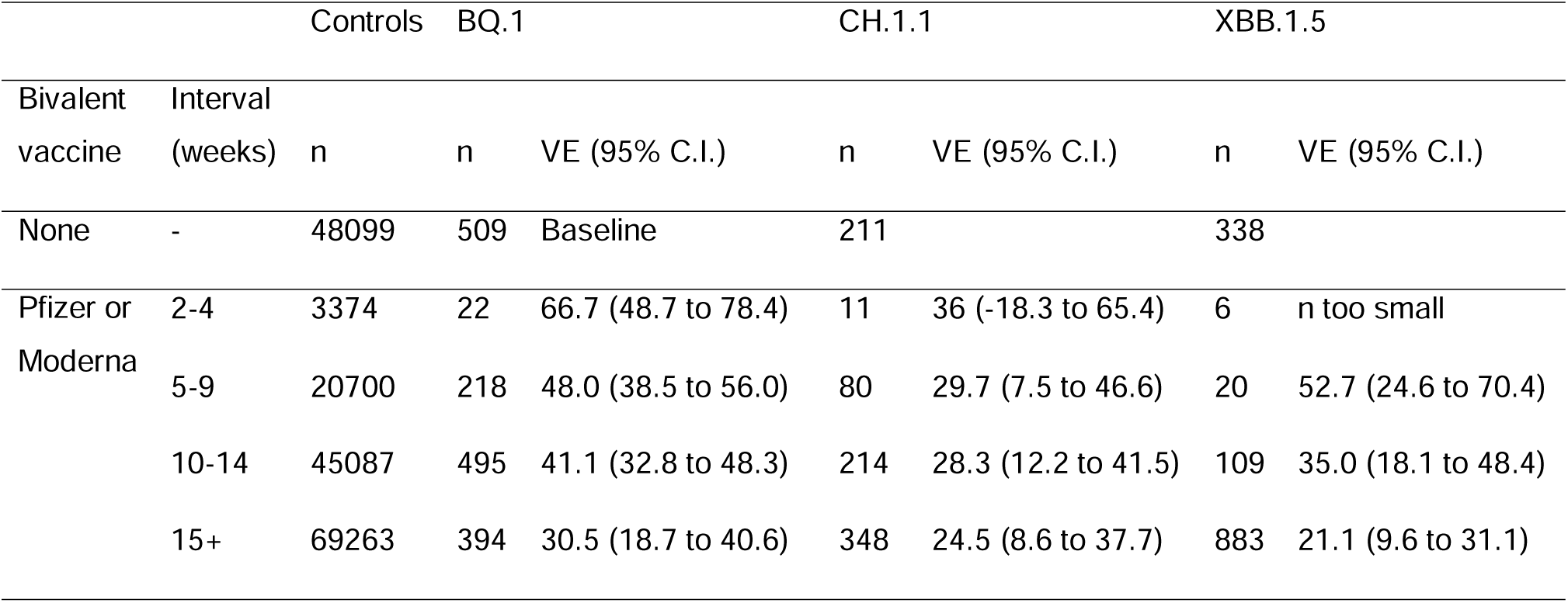
Incremental vaccine effectiveness (iVE) against hospitalisation of the bivalent BA.1 booster vaccine against BQ.1, CH.1.1 and XBB.1.5 amongst adults aged 50 years and older in England.

### Odds of admission or death after A&E attendance

4,665 individuals were identified who tested positive with BQ.1, CH.1.1 or XBB.1.5, and had a record of attendance to A&E on the same day as their positive test. Overall, the patient characteristics were broadly similar across the cohort (Supplementary Table 4). The majority of the cohort had received two or more vaccine doses, boosters, and the more recent autumn booster.

After adjusting by age group, sex, vaccination status, reinfection status, IMD quintile and geographical region, and stratifying by specimen test week, there was a significant reduction in odds after adjustment in CH.1.1 compared to BQ.1, although the upper confidence interval nears 1 (OR 0.87, 95% C.I.; 0.77 – 0.99; Table 2). However, there was no significant reduction in odds after adjustment in XBB.1.5, (OR 0.88, 95% C.I.; 0.75 – 1.02).

**Table 2.**
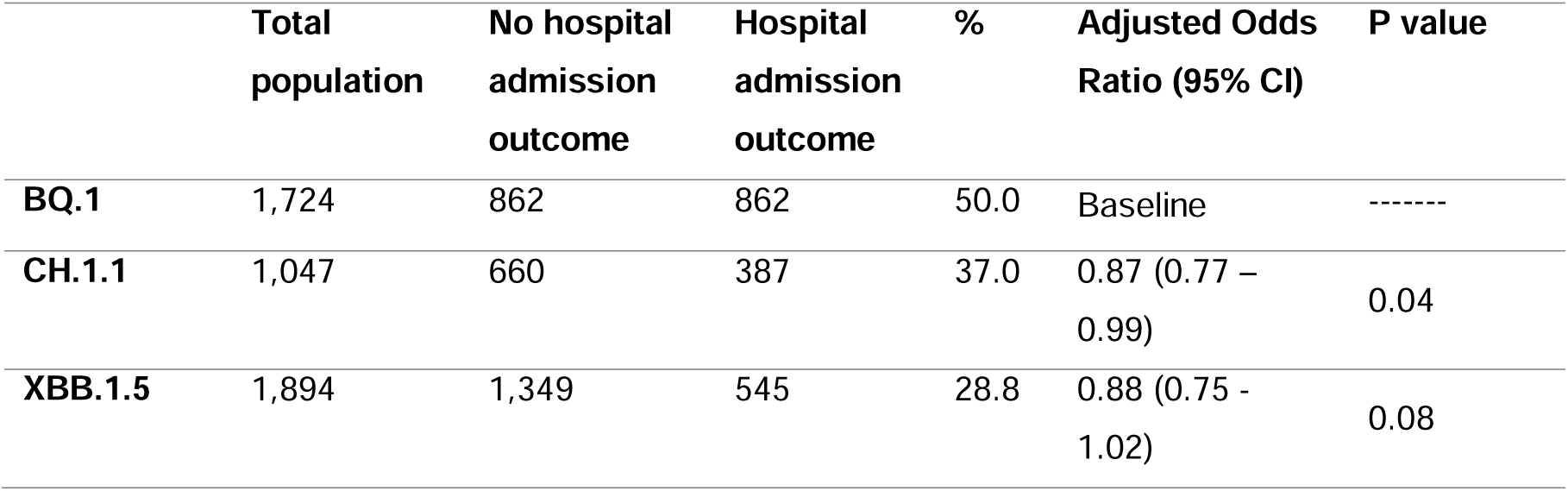
Adjusted odds ratios (OR) and 95% confidence intervals (CI) comparing risk of admission or death among individuals who attended A&E with CH.1.1 and XBB.1.5 as compared to BQ.1.

|  | **Total**<br>**population** | **No hospital**<br>**admission**<br>**outcome** | **Hospital**<br>**admission**<br>**outcome** | **%** | **Adjusted Odds**<br>**Ratio (95% CI)** | **P value** |
| --- | --- | --- | --- | --- | --- | --- |
| **BQ.1** | 1,724 | 862 | 862 | 50.0 | Baseline | ----- |
| **CH.1.1** | 1,047 | 660 | 387 | 37.0 | 0.87 (0.77 –<br>0.99) | 0.04 |
| **XBB.1.5** | 1,894 | 1,349 | 545 | 28.8 | 0.88 (0.75 -<br>1.02) | 0.08 |

### Odds of admission to ICU or death after hospitalisation

Compared to the baseline of BQ.1, there was no significant difference in the odds of admission to ICU or death for those with XBB.1.5 (OR 0.67, 95% C.I.; 0.44-1.02) or CH.1.1 (OR 0.96, 95% C.I.; 0.71–1.30) compared to BQ.1 (Table 3). Sensitivity analyses restricting to individuals with a respiratory code in their primary diagnosis field found there was a significant reduction in the odds of admission to ICU or death for those with XBB.1.5 (OR 0.48, 95% C.I.; 0.25–0.89), but not with CH.1.1 (OR 0.79, 95% C.I.; 0.53–1.17) (Supplementary Table 5, Supplementary Figure 2).

**Table 3.**
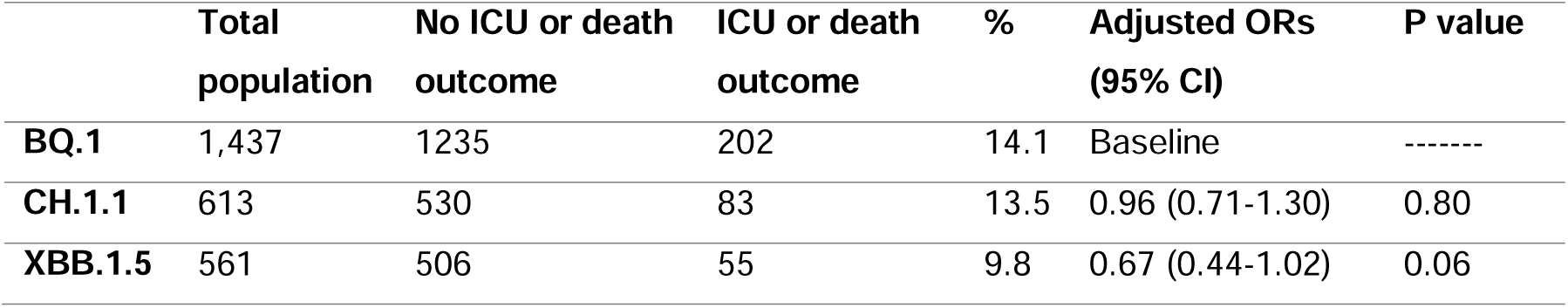
Adjusted odds ratios (OR) and 95% confidence intervals (CI) comparing risk of ICU admission or death among individuals who were admitted to hospital, and had a length of stay of two or more days, with CH.1.1 and XBB.1.5 as compared to BQ.1.

|  | **Total**<br>**population** | **No ICU or death**<br>**outcome** | **ICU or death**<br>**outcome** | **%** | **Adjusted ORs**<br>**(95% CI)** | **P value** |
| --- | --- | --- | --- | --- | --- | --- |
| **BQ.1** | 1,437 | 1235 | 202 | 14.1 | Baseline | ----- |
| **CH.1.1** | 613 | 530 | 83 | 13.5 | 0.96 (0.71-1.30) | 0.80 |
| **XBB.1.5** | 561 | 506 | 55 | 9.8 | 0.67 (0.44-1.02) | 0.06 |

### Median length of stay after hospital admission

There was no significant difference in the length of stay for those hospitalised with BQ.1 (median length of stay 5.2 days; 95% C.I.; 4.9-5.6 days), CH.1.1 (median length of stay 5.1 days; 95% C.I.; 4.6-5.6 days) or XBB.1.5 (median length of stay 5.2 days; 95% C.I.; 4.6-5.8 days) (Table 4). Sensitivity analyses restricting to those with a respiratory code in their primary diagnosis field also found no significant difference in the length of stay by variant (Supplementary Table 5).

**Table 4.** Predicted median length of stay with 95% confidence intervals (CI) of individuals who were admitted to hospital with CH.1.1 and XBB.1.5 as compared to BQ.1.

|  | **N** | **Median length of stay (95% CI) (days)** |
| --- | --- | --- |
| **BQ.1** | 1,126 | 5.2 (4.9-5.6) |
| **CH.1.1** | 518 | 5.1 (4.6-5.6) |
| **XBB.1.5** | 518 | 5.2 (4.6-5.8) |

## Discussion

We found no significant difference in the protection conferred by the bivalent BA.1 booster vaccines against hospitalisation with BQ.1, CH.1.1 and XBB.1.5. Point estimates for odds of severe disease indicators with CH.1.1 and XBB.1.5 were generally lower than for BQ.1, both for odds of hospital admission or death following A&E attendance; and for odds of ICU admission or death among hospitalised patients, though in most analyses this did not reach statistical significance. The length of stay following hospital admission also did not differ by variant.

These results follow previously observed trends, most recently with BA.4 and BA.5, showing no difference by sub-lineage in odds of admission or death following A&E attendance compared to the baseline of BA.2^8^, and similarly with BA.4.6, BA.2.75 and BQ.1 compared to BA.5^9^. Previously, we observed that VE against hospitalisation with BA.2, BA.4 and BA.5 peaked at around 60% at 2 to 14 weeks post vaccination following a third or fourth dose, estimated relative to those with waned immunity who had received their second dose at least 25 weeks prior^17^. We here found the incremental effectiveness of the vaccines in addition to at least two doses of vaccine peaked at around 48.0%, 29.7% and 52.7% for BQ.1, CH.1.1 and XBB.1.5 5 to 9 week post-vaccination. Differences in testing policy, the vaccines given and the infection histories between the study periods make it difficult to directly compare estimates, but this could indicate effectiveness is slightly reduced for current circulating sub-lineages as compared to BA.2, BA.4 and BA.5.

Since most of the adult population in England has received multiple vaccine doses and very few individuals remain unvaccinated, we considered it most relevant to estimate the additional protection the booster gave on top of that which most of the adult population eligible for an autumn booster already had^24^. A TNCC study from the US^18^ found a lower VE (around 40%) against infection with XBB/XBB.1.5 than that observed here, likely as VE is higher against more severe outcomes^16,23,28^. A study from Singapore^20^ found similar VE against XBB; in their study the VE of an mRNA vaccine (in addition to that conferred by past doses) against A&E attendance was 49% during an XBB wave.

The iVE of the bivalent boosters was comparable to that we and others have observed previously, with evidence of waning at 15 or more weeks post-vaccination^23,24,30,31^. Previously we have observed large difference in VE estimates when the hospitalisation outcome was not restricted to those with a respiratory code in the primary diagnosis field^25^, however since September 2022, PCR testing in England has been restricted to those with respiratory disease in hospital settings and in our most recent analyses we have not observed a difference in VE^22^. We therefore included all admissions regardless of ICD-10 coding in our primary VE analysis. Sensitivity analyses restricting to those with a respiratory code in the primary diagnosis field also showed no difference. We combined bivalent BA.1 booster manufacturers as we have previously not observed a difference between the Moderna BA.1 bivalent or Pfizer BA.1 bivalent vaccines^24^.

Our results do not indicate that there is a difference in the odds of an A&E presentation ending in hospital admission or death with CH.1.1 and XBB.1.5 as compared to BQ.1. Results for XBB.1.5 are consistent with previous evidence from the US on the proportion of individuals hospitalised^10^ and from Singapore in a community cohort showing no increased risk of hospitalisation^32^. The end of freely available community testing for COVID-19 in England as well as the reduction in whole genome sequencing of positive tests has provided challenges in assessing the severity of emergent variants in England. Since tests performed in hospital settings were prioritised for sequencing, we have adapted our previous methodology^33,34^ to account for this sampling bias, a limitation noted in other studies^32^, by restricting the analysis of relative severity just within individuals who attended A&E on the day of testing positive^8^.

We also found no difference in severity when assessing the odds of ICU admission or death, and by investigating the length of stay, amongst older adults. We considered the most severe outcomes were most relevant to investigate in older adults and restricted to those aged 50 and older. Sensitivity analyses restricting to those with a respiratory code in the primary diagnosis field found a decrease in the odds of ICU admission or death for XBB.1.5, as compared to BQ.1. No difference was found for CH.1.1, or for length of stay between any sub-lineage. It is possible that the decreased odds of ICU admission or death with a primary respiratory code with XBB.1.5 as compared to BQ.1 is a spurious finding due to smaller numbers of cases in the restricted analysis.

A strength of this study is the availability of real-time national-level surveillance data which has allowed us to rapidly investigate sub-lineages as they emerge. A key strength of the TNCC study design in estimating VE in contrast to a conventional cohort study or case-control design is that it helps to address unmeasured confounders related to differences in health seeking behaviours and infectious disease exposure between vaccinated and unvaccinated individuals. The TNCC requires testing to be independent of vaccination status, which is likely to be the case in a hospital setting with more severe cases. The methodology to assess the odds of hospital admission takes the approach of estimating relative severity just within cases who attended A&E. However, those attending A&E are more likely to experience severe infection than the general population. The analysis was restricted to those attending A&E on the same day as their first specimen date to account for this as those testing on the same day as presentation are more likely to represent the general population with limited access to free testing outside of healthcare settings.

Our study is an observational study that relies on hospital coding which can be prone to error. Similarly, given the observational nature of the study, there may be unmeasured confounders that we were unable to adjust for. Past infection may affect both VE and variant severity, however most infections are undocumented since freely available community testing ended. This missing data on past positivity may bias VE to be lower because past positivity is protective itself and associated with fewer vaccine doses. Not adjusting for known past positivity made little difference to estimates suggesting this is not likely to lead to a large bias. Sensitivity analyses (data not shown) demonstrated that inclusion of reinfection status had little effect on the estimates obtained on the odds of hospital admission following A&E attendance. Including those with past infection is most relevant to public health policy as most of the population have now been infected.

Together, these results provide reassuring evidence that the bivalent BA.1 booster vaccines provide similar protection against hospitalisation with BQ.1, CH.1.1 and XBB.1.5, and that both the emergent CH.1.1 and XBB.1.5 sub-lineages do not cause more severe disease as compared to BQ.1. The analyses follow previously observed trends showing similarity in the vaccine effectiveness against, and severity of, Omicron sub-lineages.

### Authors’ Contributions

JLB, NA, NAA, GD, MR conceptualised the study. NG and MC provided the genomics data. FCMK, KH, RJL, RH and JS curated the data. NG and MC provided the genomics data. FCMK, KH and RJL conducted the formal analysis, supported by NAA. FCMK, KH and RJL accessed and verified the data. FCMK and KH wrote the original draft of the manuscript. All co-authors reviewed the manuscript and were responsible for the decision to submit the manuscript.

### Funding

There was no external funding for this study.

### Ethics Committee Approval

Surveillance of COVID-19 testing and vaccination is undertaken under Regulation 3 of The Health Service (Control of Patient Information) Regulations 2002 to collect confidential patient information (www.legislation.gov.uk/uksi/2002/1438/regulation/3/made) under Sections 3(i) (a) to (c), 3(i)(d) (i) and (ii) and 3(3). The study protocol was subject to an internal review by the UK Health Security Agency Research Ethics and Governance Group and was found to be fully compliant with all regulatory requirements. As no regulatory issues were identified, and ethical review is not a requirement for this type of work, it was decided that a full ethical review would not be necessary.

### Data Sharing Statement

This work is carried out under Regulation 3 of The Health Service (Control of Patient Information; Secretary of State for Health, 2002) using patient identification information without individual patient consent as part of the UKHSA legal requirement for public health surveillance and monitoring of vaccines. As such, authors cannot make the underlying dataset publicly available for ethical and legal reasons. However, all the data used for this analysis is included as aggregated data in the manuscript tables and appendix. Applications for relevant anonymised data should be submitted to the UKHSA Office for Data Release at https://www.gov.uk/government/publications/accessing-ukhsa-protected-data.

## Data Availability

This work is carried out under Regulation 3 of The Health Service (Control of Patient Information) (Secretary of State for Health, 2002) using patient identification information without individual patient consent. Data cannot be made publicly available for ethical and legal reasons, i.e. public availability would compromise patient confidentiality as data tables list single counts of individuals rather than aggregated data.

## References

1. UK Health Security Agency. SARS-CoV-2 variants of concern and variants under investigation in England: Technical briefing 35. https://assets.publishing.service.gov.uk/government/uploads/system/uploads/attachment_data/file/1050999/Technical-Briefing-35-28January2022.pdf (2022).

2. UK Health Security Agency. SARS-CoV-2 variants of concern and variants under investigation in England: Technical briefing 36. https://assets.publishing.service.gov.uk/government/uploads/system/uploads/attachment_data/file/1054357/Technical-Briefing-36-11February2022_v2.pdf (2022).

3. UK Health Security Agency. SARS-CoV-2 variants of concern and variants under investigation in England: Technical briefing 44. https://assets.publishing.service.gov.uk/government/uploads/system/uploads/attachment_data/file/1103191/covid-technical-briefing-44-22-july-2022.pdf (2022).

4. UK Health Security Agency. SARS-CoV-2 variants of concern and variants under investigation in England: Technical briefing 48. https://assets.publishing.service.gov.uk/government/uploads/system/uploads/attachment_data/file/1120304/technical-briefing-48-25-november-2022-final.pdf (2022).

5. UK Health Security Agency. SARS-CoV-2 variants of concern and variants under investigation in England: Technical briefing 51. https://assets.publishing.service.gov.uk/government/uploads/system/uploads/attachment_data/file/1141754/variant-technical-briefing-51-10-march-2023.pdf (2023).

6. UK Health Security Agency. SARS-CoV-2 variants of concern and variants under investigation in England: Technical briefing 50. https://assets.publishing.service.gov.uk/government/uploads/system/uploads/attachment_data/file/1138007/variant-technical-briefing-50-10-february-2023.pdf (2023).

7. UK Health Security Agency. SARS-CoV-2 variants of concern and variants under investigation in England: Technical briefing 49. https://assets.publishing.service.gov.uk/government/uploads/system/uploads/attachment_data/file/1129169/variant-technical-briefing-49-11-january-2023.pdf (2023).

8. Aziz, N. A. et al. Risk of severe outcomes among SARS-CoV-2 Omicron BA.4 and BA.5 cases compared to BA.2 cases in England. Journal of Infection S0163445323002499 (2023) doi:10.1016/j.jinf.2023.04.015.

9. Seghezzo, G., et al. Risk of severe outcomes among Omicron sub-lineages BA.4.6, BA.2.75 and BQ.1 compared to BA.5 in England. http://medrxiv.org/lookup/doi/10.1101/2023.07.14.23292656 (2023) doi:10.1101/2023.07.14.23292656.

10. Luoma, E. Notes from the Field: Epidemiologic Characteristics of SARS-CoV-2 Recombinant Variant XBB.1.5 — New York City, November 1, 2022–January 4, 2023. MMWR Morb Mortal Wkly Rep 72, (2023).

11. Wang, Q. et al. Alarming antibody evasion properties of rising SARS-CoV-2 BQ and XBB subvariants. Cell 186, 279–286.e8 (2023).

12. Planas, D. et al. Resistance of Omicron subvariants BA.2.75.2, BA.4.6, and BQ.1.1 to neutralizing antibodies. Nat Commun 14, 824 (2023).

13. Kurhade, C. et al. Low neutralization of SARS-CoV-2 Omicron BA.2.75.2, BQ.1.1 and XBB.1 by parental mRNA vaccine or a BA.5 bivalent booster. Nat Med 29, 344–347 (2023).

14. Davis-Gardner, M. E. et al. Neutralization against BA.2.75.2, BQ.1.1, and XBB from mRNA Bivalent Booster. N Engl J Med 388, 183–185 (2023).

15. Uraki, R. et al. Humoral immune evasion of the omicron subvariants BQ.1.1 and XBB. The Lancet Infectious Diseases 23, 30–32 (2023).

16. Kirsebom, F. C. M. et al. COVID-19 vaccine effectiveness against the omicron (BA.2) variant in England. The Lancet Infectious Diseases 22, 931–933 (2022).

17. Kirsebom, F. C. M. et al. Effectiveness of the COVID-19 vaccines against hospitalisation with Omicron sub-lineages BA.4 and BA.5 in England. The Lancet Regional Health - Europe 23, 100537 (2022).

18. Link-Gelles, R. et al. Early Estimates of Bivalent mRNA Booster Dose Vaccine Effectiveness in Preventing Symptomatic SARS-CoV-2 Infection Attributable to Omicron BA.5- and XBB/XBB.1.5-Related Sublineages Among Immunocompetent Adults - Increasing Community Access to Testing Program, United States, December 2022-January 2023. MMWR Morb Mortal Wkly Rep 72, 119–124 (2023).

19. Lin, D.-Y. et al. Durability of Bivalent Boosters against Omicron Subvariants. N Engl J Med 388, 1818–1820 (2023).

20. Wee, L. E. et al. Long-term Real-world Protection Afforded by Third mRNA Doses Against Symptomatic SARS-COV-2 Infections, COVID-19-related Emergency Attendances and Hospitalizations Amongst Older Singaporeans During an Omicron XBB Wave. Clinical Infectious Diseases ciad345 (2023) doi:10.1093/cid/ciad345.

21. UK Health Security Agency. COVID-19: the green book, chapter 14a. Immunisation against infectious diseases. (2020).

22. Joint Committee on Vaccination and Immunisation. JCVI updated statement on the COVID-19 vaccination programme for autumn 2022. https://www.gov.uk/government/publications/jcvi-updated-statement-on-the-covid-19-vaccination-programme-for-autumn-2022 (2022).

23. Tenforde, M. et al. Early Estimates of Bivalent mRNA Vaccine Effectiveness in Preventing COVID-19–Associated Emergency Department or Urgent Care Encounters and Hospitalizations Among Immunocompetent Adults — VISION *Network, Nine States, September–November 2022*. https://www.cdc.gov/mmwr/volumes/71/wr/mm7153a1.htm?s_cid=mm7153a1_w (2023).

24. Kirsebom, F. C. M., Andrews, N., Stowe, J., Ramsay, M. & Bernal, J.L. Duration of protection of ancestral-strain monovalent vaccines and effectiveness of bivalent BA.1 boosters against COVID-19 hospitalisation in England: a test-negative case-control study. The Lancet Infectious Diseases 0, 0 (2023).

25. Stowe, J., Andrews, N., Kirsebom, F., Ramsay, M. & Bernal, J. L. Effectiveness of COVID-19 vaccines against Omicron and Delta hospitalisation, a test negative case-control study. Nat Commun 13, 5736 (2022).

26. Andrews, N. et al. Covid-19 Vaccine Effectiveness against the Omicron (B.1.1.529) Variant. N Engl J Med 386, 1532–1546 (2022).

27. UKHSA Genomics Public Health Analysis. UKHSA Standardised Variant Definitions. https://github.com/ukhsa-collaboration/variant_definitions.

28. NHS England. National Vaccination Programmes. NHS England https://www.england.nhs.uk/contact-us/privacy-notice/national-flu-vaccination-programme/#immunisation.

29. Bhattacharya, A. et al. Healthcare-associated COVID-19 in England: A national data linkage study. J Infect 83, 565–572 (2021).

30. Lin, D.-Y. et al. Effectiveness of Bivalent Boosters against Severe Omicron Infection. N Engl J Med 388, 764–766 (2023).

31. Surie, D. et al. Early Estimates of Bivalent mRNA Vaccine Effectiveness in Preventing COVID-19-Associated Hospitalization Among Immunocompetent Adults Aged ≥65 Years - IVY Network, 18 States, September 8-November 30, 2022. MMWR Morb Mortal Wkly Rep 71, 1625–1630 (2022).

32. Pung, R., et al. Severity of SARS-CoV-2 Omicron XBB subvariants in Singapore. http://medrxiv.org/lookup/doi/10.1101/2023.05.04.23289510 (2023) doi:10.1101/2023.05.04.23289510.

33. Webster, H. H. et al. Hospitalisation and mortality risk of SARS-COV-2 variant omicron sub-lineage BA.2 compared to BA.1 in England. Nat Commun 13, 6053 (2022).

34. Nyberg, T. et al. Comparative analysis of the risks of hospitalisation and death associated with SARS-CoV-2 omicron (B.1.1.529) and delta (B.1.617.2) variants in England: a cohort study. The Lancet 399, 1303–1312 (2022).

