## Supplementary Appendix for "Vaccine effectiveness against hospitalisation and comparative odds of hospital admission and severe outcomes with BQ.1, CH.1.1. and XBB.1.5 in England"

### Data Sources

#### COVID-19 Testing Data

Polymerase-chain-reaction (PCR) testing for SARS CoV-2 in the United Kingdom is undertaken by hospital and public health laboratories (Pillar 1). Prior to 1^st^ April 2022, community testing (Pillar 2) was available with the use of drive through or at-home testing to anyone with symptoms consistent with COVID-19 (high temperature, new continuous cough, or loss or change in sense of smell or taste), is a contact of a confirmed case, for care home staff and residents or who has self-tested as positive using a lateral flow device. Since the end of freely available community testing on 1^st^ April 2022, most PCR testing in England has been done in hospital settings and since September 2022, Pillar 1 (hospital) PCR testing has been restricted to those with respiratory disease in hospital settings.

Negative tests taken within 7 days of a previous negative test were dropped as these likely represent the same episode. Negative tests taken within 21 days of a subsequent positive test were also excluded as chances are high that these are false negatives. Tests within 90 days of a previous positive test were also excluded as these likely represent the same episode. The date of an individual’s most recent prior positive test was identified from all historic testing data (Pillar 1 or Pillar 2). For the VE analyses, individuals contributed a maximum of one negative control test (selected at random) in each period of 13^th^ June to 4^th^ September 2022, and 5^th^ September 2022 onwards. Variant status was classified based on whole genome sequencing. Any non-Omicron cases were excluded. Further details on the reasons for these exclusions are described previously^1^.

#### National Immunisation Management System (NIMS)

The National Immunisation Management System (NIMS) is a national vaccine register containing demographic information on the whole population of England registered with a GP. It is used to record all COVID-19 vaccinations. Testing data were linked to NIMS using combinations of the unique individual NHS number, date of birth, surname, first name, and postcode using deterministic linkage. NIMS was accessed for dates of vaccination and manufacturer, sex, date of birth, ethnicity, and residential address. Addresses were used to determine index of multiple deprivation quintile. Data on risk group status (those identified as at risk previously in the pandemic and those identified recently as requiring an autumn booster by NHS CaaS (Cohorting as a Service^2^), clinically extremely vulnerable status, severely immunosuppressed and health/social care worker status were also extracted from the NIMS. Bivalent doses given as part of the autumn booster programme were classified based on SNOMED coding and timing (administered after 5^th^ September 2022).

Where vaccination status was adjusted for, vaccination status was categorised based on receipt of the dose or booster at least two weeks prior to positive specimen date.

#### Emergency Care Hospital Data

Emergency Care hospital attendances from the Emergency Care Dataset (ECDS), which includes hospital attendances through emergency departments but not elective admissions, were linked to cases to identify attendances on the same day as their first positive PCR test. Admissions were identified where the record discharge was either an admission or transfer.

#### Hospital Admission Data

Secondary Uses Service (SUS) is the national electronic database of hospital admissions that provides timely updates of ICD-10 codes for completed hospital stays for all NHS hospitals in England^3^. Up to 24 ICD-10 diagnoses fields can be completed in SUS for each admission with the first diagnosis field indicating the primary reason for admission. Oxygen use and ventilation support was ascertained using the Classification of Interventions and Procedures (OPCS-4) codes. Intensive Care Unit (ICU) admission status was ascertained by the Main Specialty of the ward being Critical Care Medicine or the Treatment Function being Intensive Care Medicine. Length of stay was calculated as date of discharge minus the date of admission.

#### Mortality data

Mortality was obtained from the UKHSA COVID-19 mortality dataset^4^ or from the National Immunisation Management System where it was identified from the GP record. Deaths were only included where the date of death was within 28 days of earliest specimen date.

### Supplementary Tables and Figures

Supplementary Figure 1. Prevalence of SARS-CoV-2 variants amongst available sequences episodes for England from 18 April 2022 up to 2 April 2023. The grey line indicates proportion of cases sequenced. The vertical dashed lines (red) denote changes in policies: April 2022 denotes the start of England’s ‘Living with COVID’ Plan. End of August line denotes the changes in asymptomatic testing. April 2023 line denotes further changes in testing policy. Note: Recombinants such as XD, are not specified but are largely within the ‘Other’ group currently as numbers are too small.

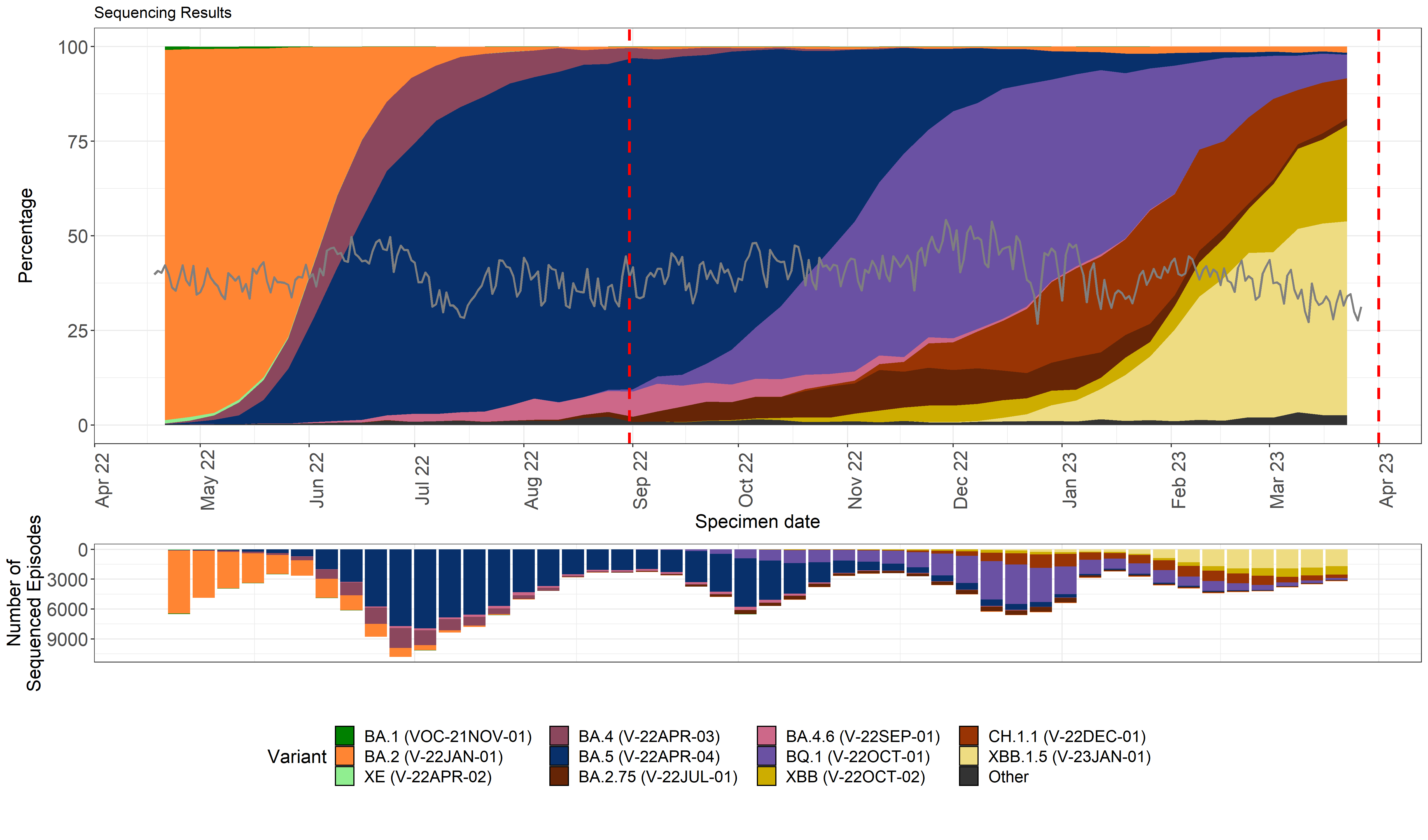

Supplementary Table 1. SUS Acute respiratory illness ICD-10 code list.

| SUS Acute respiratory illness ICD10 code list | |
| --- | --- |
| J04* | Acute laryngitis and tracheitis |
| J09* | Influenza due to identified avian influenza virus |
| J10* | Influenza with pneumonia, other influenza virus identified |
| J11* | Influenza with pneumonia, virus not identified |
| J12* | Viral pneumonia, not elsewhere classified |
| J13* | Pneumonia due to Streptococcus pneumoniae |
| J14* | Pneumonia due to Haemophilus influenzae |
| J15* | Bacterial pneumonia, not elsewhere classified |
| J16* | Pneumonia due to other infectious organisms, not elsewhere classified |
| J17* | Pneumonia in diseases classified elsewhere |
| J18* | Pneumonia, organism unspecified |
| J20* | Acute bronchitis |
| J21* | Acute bronchiolitis |
| J22* | Unspecified acute lower respiratory infection |
| J80* | ARDS (related to respiratory infection) |
| U071, U072 | COVID-19, virus identified and not identified |
| U04* | Severe acute respiratory syndrome (SARS) |

Supplementary Table 2. Descriptive characteristics of those included the incremental vaccine effectiveness (iVE) analysis of the bivalent BA.1 booster vaccines.

|  |  |  | Overall | | Controls | | BQ.1 | | CH.1.1 | | XBB.1.5 | |
| --- | --- | --- | --- | --- | --- | --- | --- | --- | --- | --- | --- | --- |
|  |  |  | n | % | n | % | n | % | n | % | n | % |
| Vaccination Status | Bivalent vaccine | Interval | 191,229 | 100% | 187,348 | 98% | 1,647 | 1% | 877 | 0% | 1,357 | 82% |
|  | None |  | 49,167 | 25.7% | 48,099 | 25.7% | 509 | 30.9% | 221 | 25.2% | 338 | 24.9% |
|  | Pfizer or Moderna | 0-6 days | 257 | 0.1% | 251 | 0.1% | 5 | 0.3% | 1 | 0.1% | 0 | 0.0% |
|  |  | 7-13 days | 581 | 0.3% | 574 | 0.3% | 4 | 0.2% | 2 | 0.2% | 1 | 0.1% |
|  |  | 2-4 weeks | 3,413 | 1.8% | 3,374 | 1.8% | 22 | 1.3% | 11 | 1.3% | 6 | 0.4% |
|  |  | 5-9 weeks | 21,018 | 11.0% | 20,700 | 11.0% | 218 | 13.2% | 80 | 9.1% | 20 | 1.5% |
|  |  | 10-14 weeks | 45,905 | 24.0% | 45,087 | 24.1% | 495 | 30.1% | 214 | 24.4% | 109 | 8.0% |
|  |  | 15+ weeks | 70,888 | 37.1% | 69,263 | 37.0% | 394 | 23.9% | 348 | 39.7% | 883 | 65.1% |
| Gender | Female |  | 96,303 | 50.4% | 94,471 | 50.4% | 769 | 46.7% | 435 | 49.6% | 628 | 46.3% |
|  | Male |  | 92,656 | 48.5% | 90,608 | 48.4% | 878 | 53.3% | 442 | 50.4% | 728 | 53.6% |
|  | Missing |  | 2270 | 1.2% | 2,269 | 1.2% | 0 | 0.0% | 0 | 0.0% | 1 | 0.1% |
| Age | 50-54 |  | 21,526 | 11.3% | 10,671 | 5.7% | 40 | 2.4% | 33 | 3.8% | 10,782 | 794.5% |
|  | 55-59 |  | 28,849 | 15.1% | 14,309 | 7.6% | 58 | 3.5% | 29 | 3.3% | 14,453 | 1065.1% |
|  | 60-64 |  | 34,923 | 18.3% | 17,283 | 9.2% | 92 | 5.6% | 46 | 5.2% | 17,502 | 1289.8% |
|  | 65-69 |  | 40,945 | 21.4% | 20,231 | 10.8% | 128 | 7.8% | 69 | 7.9% | 20,517 | 1511.9% |
|  | 70-74 |  | 51,746 | 27.1% | 25,553 | 13.6% | 163 | 9.9% | 81 | 9.2% | 25,949 | 1912.2% |
|  | 75-79 |  | 61,403 | 32.1% | 30,158 | 16.1% | 277 | 16.8% | 154 | 17.6% | 30,814 | 2270.7% |
|  | 80-84 |  | 56,662 | 29.6% | 27,730 | 14.8% | 313 | 19.0% | 159 | 18.1% | 28,460 | 2097.3% |
|  | 85-89 |  | 49,445 | 25.9% | 24,096 | 12.9% | 322 | 19.6% | 172 | 19.6% | 24,855 | 1831.6% |
|  | 90+ |  | 35,602 | 18.6% | 17,317 | 9.2% | 254 | 15.4% | 134 | 15.3% | 17,897 | 1318.9% |
| Ethnicity | African |  | 1289 | 0.7% | 1,272 | 0.7% | 7 | 0.4% | 1 | 0.1% | 9 | 0.7% |
|  | Any other Asian background | | 1683 | 0.9% | 1,659 | 0.9% | 12 | 0.7% | 2 | 0.2% | 10 | 0.7% |
|  | Any other Black background | | 573 | 0.3% | 567 | 0.3% | 4 | 0.2% | 1 | 0.1% | 1 | 0.1% |
|  | Any other White background | | 8,831 | 4.6% | 8,642 | 4.6% | 75 | 4.6% | 35 | 4.0% | 79 | 5.8% |
|  | Any other ethnic group | | 1793 | 0.9% | 1,759 | 0.9% | 7 | 0.4% | 8 | 0.9% | 19 | 1.4% |
|  | Any other mixed background | | 657 | 0.3% | 645 | 0.3% | 5 | 0.3% | 1 | 0.1% | 6 | 0.4% |
|  | Bangladeshi or British Bangladeshi | | 757 | 0.4% | 750 | 0.4% | 1 | 0.1% | 2 | 0.2% | 4 | 0.3% |
|  | British, Mixed British | | 157,227 | 82.2% | 153,937 | 82.2% | 1,421 | 86.3% | 749 | 85.4% | 1,120 | 82.5% |
|  | Caribbean |  | 1504 | 0.8% | 1,483 | 0.8% | 9 | 0.5% | 6 | 0.7% | 6 | 0.4% |
|  | Chinese |  | 364 | 0.2% | 360 | 0.2% | 0 | 0.0% | 0 | 0.0% | 4 | 0.3% |
|  | Indian or British Indian | | 4310 | 2.3% | 4,266 | 2.3% | 15 | 0.9% | 18 | 2.1% | 11 | 0.8% |
|  | Irish |  | 2223 | 1.2% | 2,178 | 1.2% | 17 | 1.0% | 12 | 1.4% | 16 | 1.2% |
|  | Pakistani or British Pakistani | | 2193 | 1.1% | 2,174 | 1.2% | 7 | 0.4% | 4 | 0.5% | 8 | 0.6% |
|  | White and Asian | | 189 | 0.1% | 187 | 0.1% | 0 | 0.0% | 2 | 0.2% | 0 | 0.0% |
|  | White and Black African | | 168 | 0.1% | 163 | 0.1% | 4 | 0.2% | 0 | 0.0% | 1 | 0.1% |
|  | White and Black Caribbean | | 293 | 0.2% | 288 | 0.2% | 3 | 0.2% | 0 | 0.0% | 2 | 0.1% |
|  | Missing |  | 7,175 | 3.8% | 7,018 | 3.7% | 60 | 3.6% | 36 | 4.1% | 61 | 4.5% |
| NHS Region | East of England | | 15,260 | 8.0% | 14,841 | 7.9% | 165 | 10.0% | 95 | 10.8% | 159 | 11.7% |
|  | London |  | 25,916 | 13.6% | 25,533 | 13.6% | 167 | 10.1% | 60 | 6.8% | 156 | 11.5% |
|  | Midlands |  | 41,561 | 21.7% | 40,788 | 21.8% | 351 | 21.3% | 165 | 18.8% | 257 | 18.9% |
|  | North East |  | 38,197 | 20.0% | 37,568 | 20.1% | 262 | 15.9% | 132 | 15.1% | 235 | 17.3% |
|  | North West |  | 23,082 | 12.1% | 22,627 | 12.1% | 208 | 12.6% | 71 | 8.1% | 176 | 13.0% |
|  | South East |  | 25,483 | 13.3% | 24,812 | 13.2% | 279 | 16.9% | 140 | 16.0% | 252 | 18.6% |
|  | South West |  | 21,730 | 11.4% | 21,179 | 11.3% | 215 | 13.1% | 214 | 24.4% | 122 | 9.0% |
| IMD Quintiles | 1 |  | 39,550 | 20.7% | 38,786 | 20.7% | 334 | 20.3% | 151 | 17.2% | 279 | 20.6% |
|  | 2 |  | 38,429 | 20.1% | 37,682 | 20.1% | 333 | 20.2% | 178 | 20.3% | 236 | 17.4% |
|  | 3 |  | 38,604 | 20.2% | 37,815 | 20.2% | 313 | 19.0% | 189 | 21.6% | 287 | 21.1% |
|  | 4 |  | 38,515 | 20.1% | 37,717 | 20.1% | 347 | 21.1% | 178 | 20.3% | 273 | 20.1% |
|  | 5 |  | 35,486 | 18.6% | 34,720 | 18.5% | 318 | 19.3% | 175 | 20.0% | 273 | 20.1% |
|  | Missing |  | 645 | 0.3% | 628 | 0.3% | 2 | 0.1% | 6 | 0.7% | 9 | 0.7% |
| Previously positive | None |  | 144,371 | 75.5% | 141,013 | 75.3% | 1,430 | 86.8% | 752 | 85.7% | 1,176 | 86.7% |
|  | Wild-type |  | 4,009 | 2.1% | 3,951 | 2.1% | 25 | 1.5% | 15 | 1.7% | 18 | 1.3% |
|  | Alpha |  | 5,304 | 2.8% | 5,239 | 2.8% | 31 | 1.9% | 12 | 1.4% | 22 | 1.6% |
|  | Delta |  | 6,163 | 3.2% | 6,069 | 3.2% | 40 | 2.4% | 27 | 3.1% | 27 | 2.0% |
|  | Omicron (before 1st April 2022) | | 15,606 | 8.2% | 15,411 | 8.2% | 87 | 5.3% | 44 | 5.0% | 64 | 4.7% |
|  | Omicron (from 1st April 2022 onwards) | | 15,776 | 8.2% | 15,665 | 8.4% | 34 | 2.1% | 27 | 3.1% | 50 | 3.7% |
| Risk status | Healthcare worker | | 1,835 | 1.0% | 1,821 | 1.0% | 7 | 0.4% | 4 | 0.5% | 3 | 0.2% |
|  | Carehome |  | 5,792 | 3.0% | 5,619 | 3.0% | 62 | 3.8% | 45 | 5.1% | 66 | 4.9% |
|  | CaaS autumn booster cohort | | 169,872 | 88.8% | 166,249 | 88.7% | 1,554 | 94.4% | 824 | 94.0% | 1,245 | 91.7% |
|  | At risk - ever, from NIMS* | | 35,521 | 18.6% | 35,093 | 18.7% | 165 | 10.0% | 100 | 11.4% | 163 | 12.0% |
|  | CEV |  | 70,811 | 37.0% | 69,157 | 36.9% | 680 | 41.3% | 382 | 43.6% | 592 | 43.6% |
|  | Severely immunosuppressed | | 17,274 | 9.0% | 16,844 | 9.0% | 182 | 11.1% | 98 | 11.2% | 150 | 11.1% |
| *Only for those <65 years | |  |  |  |  |  |  |  |  |  |  |  |

Supplementary Figure 2. Proportion of BQ.1, CH.1.1 and XBB.1.5 variant cases over time in the study period.

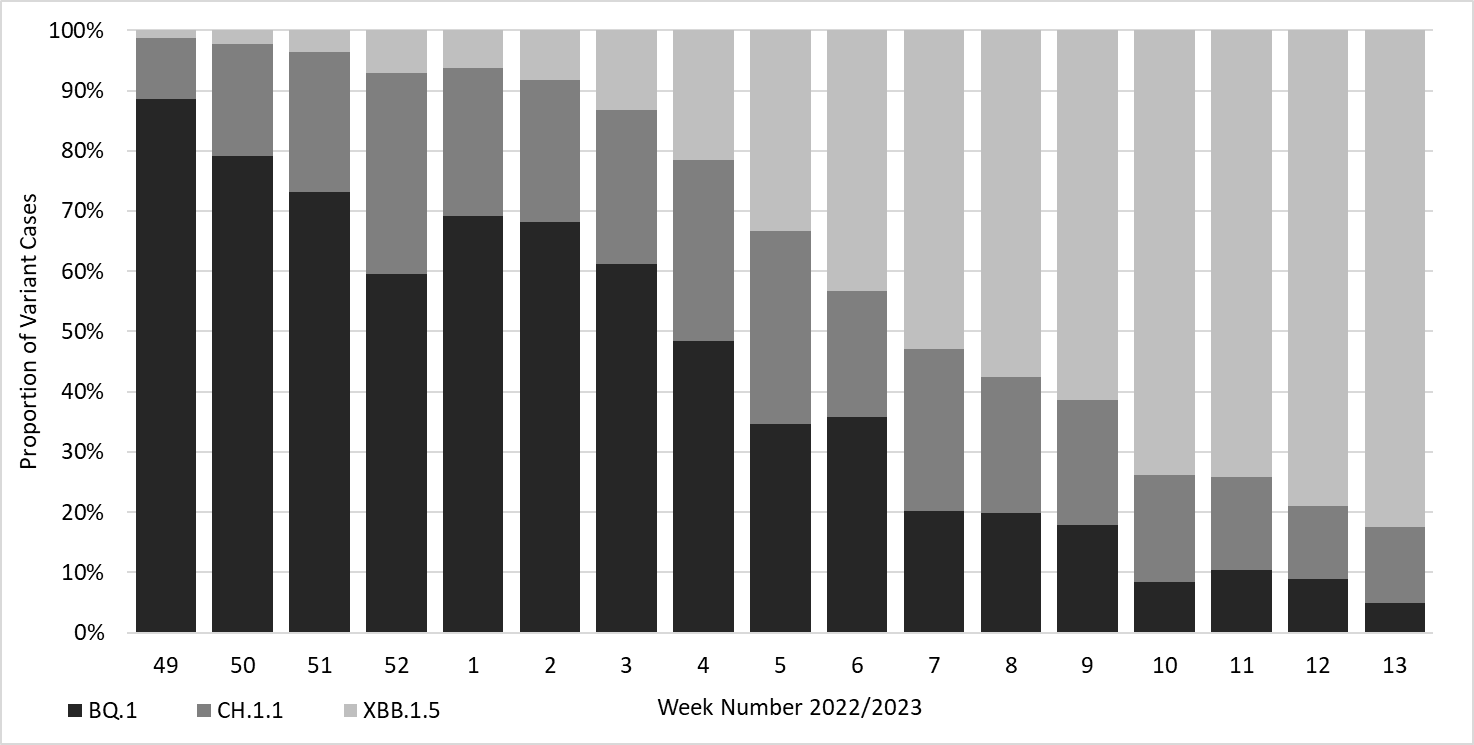

Supplementary Table 3. Incremental vaccine effectiveness (iVE) against hospitalisation of the bivalent BA.1 booster vaccine against BQ.1, CH.1.1 and XBB.1.5 amongst adults aged 50 years and older in England, restricted to those with an ARI code in the primary diagnosis field.

|  |  | Controls | BQ.1 | | CH.1.1 | | XBB.1.5 | |
| --- | --- | --- | --- | --- | --- | --- | --- | --- |
| Bivalent vaccine | Interval (weeks) | n | n | VE (95% C.I.) | n | VE (95% C.I.) | n | VE (95% C.I.) |
| None | - | 9582 | 257 |  | 99 |  | 111 |  |
| Pfizer or Moderna | 2-4 | 875 | 13 | 60.7 (30.7 to 77.7) | 8 | 2 (-104.8 to 53.1) | 4 | n too small |
|  | 5-9 | 5636 | 115 | 46.6 (32.7 to 57.7) | 41 | 27 (-7.4 to 50.3) | 9 | 42 (-18 to 71.5) |
|  | 10-14 | 12544 | 274 | 39.8 (27.7 to 49.8) | 115 | 28.4 (4.4 to 46.4) | 53 | 21.8 (-11.7 to 45.2) |
|  | 15+ | 9758 | 165 | 30.9 (12.6 to 45.4) | 125 | 32.3 (8.5 to 49.9) | 292 | 15.8 (-6.8 to 33.7) |

Supplementary Table 4. Descriptive characteristics of those included in the risk of admission or death after A&E attendance analysis.

|  | BQ.1 (n = 1,724) | | CH.1.1 (n = 1,047) | | XBB.1.5 (n = 1,894) | |
| --- | --- | --- | --- | --- | --- | --- |
|  | Cases (%)  (n = 862) | Controls (%)  (n = 862) | Cases (%)  (n = 387) | Controls (%)  (n = 660) | Cases (%)  (n = 545) | Controls (%)  (n = 1,349) |
| *Age group* |  |  |  |  |  |  |
| 0-9 | 28 (3.2) | 105 (12.2) | 17 (4.4) | 71 (10.8) | 25 (4.6) | 142 (10.5) |
| 10-19 | 5 (0.6) | 15 (1.7) | 2 (0.5) | 19 (2.9) | 10 (1.8) | 24 (1.8) |
| 20-29 | 4 (0.5) | 43 (5.0) | 7 (1.8) | 38 (5.8) | 10 (1.8) | 76 (5.6) |
| 30-39 | 20 (2.3) | 60 (7.0) | 8 (2.1) | 42 (6.4) | 15 (2.8) | 83 (6.2) |
| 40-49 | 17 (2.0) | 47 (5.5) | 5 (1.3) | 41 (6.2) | 15 (2.8) | 67 (5.0) |
| 50-59 | 45 (5.2) | 73 (8.5) | 29 (7.5) | 58 (8.8) | 30 (5.5) | 108 (8.0) |
| 60-69 | 98 (11.4) | 93 (10.8) | 58 (15.0) | 80 (12.1) | 56 (10.3) | 129 (9.6) |
| 70-79 | 222 (25.8) | 158 (18.3) | 84 (21.7) | 119 (18.0) | 131 (24.0) | 236 (17.5) |
| 80-89 | 298 (34.6) | 199 (23.1) | 130 (33.6) | 135 (20.5) | 170 (31.2) | 355 (26.3) |
| ≥ 90 | 125 (14.5) | 69 (8.0) | 47 (12.1) | 57 (8.6) | 83 (15.2) | 129 (9.6) |
| *Sex* |  |  |  |  |  |  |
| Female | 407 (47.2) | 430 (49.9) | 204 (52.7) | 337 (51.1) | 273 (50.1) | 659 (48.9) |
| Male | 455 (52.8) | 432 (50.1) | 183 (47.3) | 323 (48.9) | 272 (49.9) | 690 (51.1) |
| *Vaccination status* |  |  |  |  |  |  |
| Unvaccinated | 83 (9.6) | 190 (22.0) | 45 (11.6) | 112 (17.0) | 59 (10.8) | 245 (18.2) |
| Dose 1 or 2 | 70 (8.1) | 97 (11.3) | 28 (7.2) | 77 (11.7) | 34 (6.2) | 152 (11.3) |
| Dose 2/3 + early and/or spring booster | 223 (25.9) | 177 (20.5) | 79 (20.4) | 133 (20.2) | 118 (21.7) | 245 (18.2) |
| Dose 2/3 + autumn booster (inc. other boosters) | 486 (56.4) | 398 (46.2) | 235 (60.7) | 338 (51.2) | 334 (61.3) | 707 (52.4) |
| *Prior infection status* |  |  |  |  |  |  |
| First infection | 734 (85.2) | 743 (86.2) | 333 (86.0) | 552 (83.6) | 452 (82.9) | 1,129 (83.7) |
| Reinfection | 128 (14.8) | 119 (13.8) | 54 (14.0) | 108 (16.4) | 93 (17.1) | 220 (16.3) |
| *Index of multiple deprivation quintile* |  |  |  |  |  |  |
| 1 (Most deprived) | 209 (24.2) | 177 (20.5) | 84 (21.7) | 111 (16.8) | 141 (25.9) | 294 (21.8) |
| 2 | 167 (19.4) | 201 (23.3) | 74 (19.1) | 135 (20.5) | 100 (18.3) | 273 (20.2) |
| 3 | 178 (20.6) | 159 (18.4) | 78 (20.2) | 132 (20.0) | 119 (21.8) | 266 (19.7) |
| 4 | 162 (18.8) | 168 (19.5) | 75 (19.4) | 143 (21.7) | 93 (17.1) | 265 (19.6) |
| 5 (Least deprived) | 146 (16.9) | 157 (18.2) | 76 (19.6) | 139 (21.1) | 92 (16.9) | 251 (18.6) |
| *Region* |  |  |  |  |  |  |
| London | 131 (15.2) | 135 (15.7) | 46 (11.9) | 72 (10.9) | 100 (18.3) | 208 (15.4) |
| Midlands | 139 (16.1) | 171 (19.8) | 68 (17.6) | 120 (18.2) | 96 (17.6) | 249 (18.5) |
| East of England | 91 (10.6) | 82 (9.5) | 35 (9.0) | 68 (10.3) | 43 (7.9) | 157 (11.6) |
| North East and Yorkshire | 150 (17.4) | 90 (10.4) | 58 (15.0) | 72 (10.9) | 117 (21.5) | 175 (13.0) |
| North West | 103 (11.9) | 103 (11.9) | 28 (7.2) | 67 (10.2) | 82 (15.0) | 150 (11.1) |
| South East | 144 (16.7) | 170 (19.7) | 50 (12.9) | 107 (16.2) | 57 (10.5) | 238 (17.6) |
| South West | 104 (12.1) | 111 (12.9) | 102 (26.4) | 154 (23.3) | 50 (9.2) | 172 (12.8) |

Supplementary Table 5. Adjusted odds ratios (OR) and 95% confidence intervals (CI) comparing odds of ICU admission or death among individuals who were admitted to hospital, had a length of stay of two or more days and a respiratory code in their primary diagnosis field, for cases with Omicron CH.1.1 and XBB.1.5 compared to Omicron BQ.1. * denotes a significant result (p<0.05).

|  | Total population | No ICU or death outcome | ICU or death outcome | % | Adjusted OR  (95% CI) | P value |
| --- | --- | --- | --- | --- | --- | --- |
| BQ.1 | 763 | 631 | 132 | 17.3 | Baseline | ------- |
| CH.1.1 | 316 | 271 | 45 | 14.2 | 0.79 (0.53-1.17) | 0.239 |
| XBB.1.5 | 206 | 184 | 22 | 10.7 | 0.48 (0.25-0.89) | 0.021* |

Supplementary Figure 3. Adjusted odds ratios (OR) and 95% confidence intervals (CI) comparing odds of ICU admission or death among individuals who were admitted to hospital, had a length of stay of two or more days and a respiratory code in their primary diagnosis field, with CH.1.1 and XBB.1.5 as compared to BQ.1.

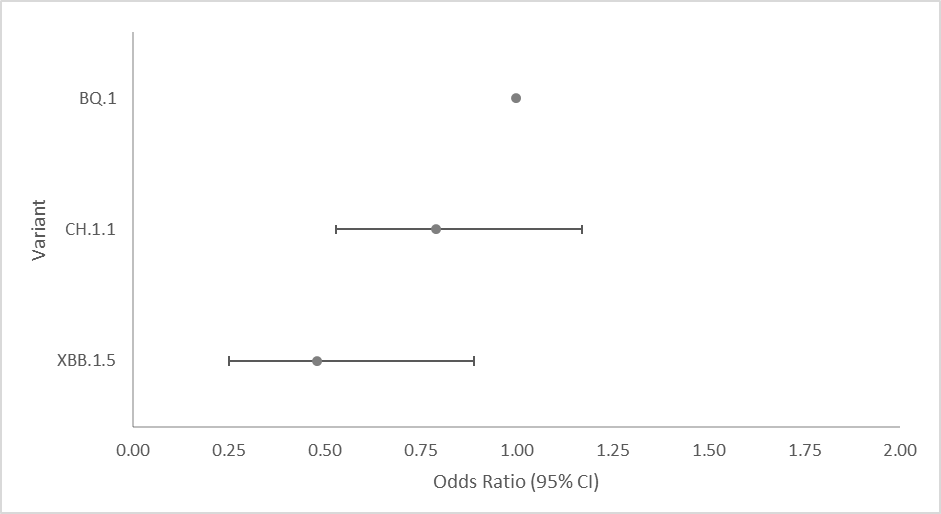

Odds Ratio (95% CI)

| Variant | HR |
| --- | --- |
| XBB.1.5 | 0.48 |
| CH.1.1 | 0.79 |
| BQ.1 | 1 |

|  |  |
| --- | --- |
| 0.48 | 0.5 |
| 0.79 | 1.5 |
| 1 | 2.5 |

Supplementary Table 5. Median length of stay with 95% confidence intervals (CI) of individuals who were admitted to hospital and had a respiratory code in the primary diagnosis field, with CH.1.1 and XBB.1.5 as compared to BQ.1.

|  | N | Median length of stay (95% CI) (days) |
| --- | --- | --- |
| BQ.1 | 571 | 5.1 (4.6-5.6) |
| CH.1.1 | 253 | 4.8 (4.2-5.5) |
| XBB.1.5 | 207 | 4.4 (3.6-5.2) |

### References

1.	Andrews, N. *et al.* Covid-19 Vaccine Effectiveness against the Omicron (B.1.1.529) Variant. *N. Engl. J. Med.* **386**, 1532–1546 (2022).

2.	NHS Digital. Cohorting as a Service (CaaS). https://digital.nhs.uk/services/cohorting-as-a-service-caas (2023).

3.	NHS Digital. Secondary Uses Service (SUS). https://digital.nhs.uk/services/secondary-uses-service-sus (2023).

4.	Brown, A. E. *et al.* Epidemiology of Confirmed COVID-19 Deaths in Adults, England, March-December 2020. *Emerg. Infect. Dis.* **27**, 1468–1471 (2021).
